# Investigation of Autosomal Genetic Sex Differences in Parkinson’s disease

**DOI:** 10.1101/2021.02.09.21250262

**Authors:** Cornelis Blauwendraat, Hirotaka Iwaki, Mary B. Makarious, Sara Bandres-Ciga, Hampton Leonard, Francis P. Grenn, Julie Lake, Lynne Krohn, Manuela Tan, Jonggeol Jeff Kim, Jesse Raphael Gibbs, Dena G. Hernandez, Jennifer A. Ruskey, Lasse Pihlstrøm, Mathias Toft, Jacobus J. van Hilten, Johan Marinus, Claudia Schulte, Kathrin Brockmann, Manu Sharma, Ari Siitonen, Kari Majamaa, Johanna Eerola-Rautio, Pentti J. Tienari, Donald Grosset, Suzanne Lesage, Jean-Christophe Corvol, Alexis Brice, Nick Wood, John Hardy, Ziv Gan-Or, Peter Heutink, Thomas Gasser, Huw Morris, Alastair J. Noyce, Mike A. Nalls, Andrew B. Singleton, on behalf of the International Parkinson’s Disease Genomics Consortium (IPDGC)

**Author notes:** **Correspondence to:** Cornelis Blauwendraat, Laboratory of Neurogenetics, NIA, NIH, Building 35, 35 Convent Drive, Bethesda, MD 20892, USA.

## Abstract

Parkinson’s disease (PD) is a complex neurodegenerative disorder. Males are on average ∼1.5 times more likely to develop PD compared to females. Over the years genome-wide association studies (GWAS) have identified numerous genetic risk factors for PD, however it is unclear whether genetics contribute to disease etiology in a sex-specific manner.

In an effort to study sex-specific genetic factors associated with PD, we explored two large genetic datasets from the International Parkinson’s Disease Genomics Consortium and the UK Biobank consisting of 13,020 male PD cases, 7,936 paternal proxy cases, 89,660 male controls, 7,947 female PD cases, 5,473 maternal proxy cases and 90,662 female controls. We performed GWAS meta-analyses to identify distinct patterns of genetic risk contributing to disease in male versus female PD cases.

In total 19 genome-wide significant regions were identified, and no sex-specific effects were observed. A high genetic correlation between the male and female PD GWASes was identified (rg=0.877) and heritability estimates were identical between male and female PD cases (∼20%).

We did not detect any significant genetic differences between male or female PD cases. Our study does not support the notion that common genetic variation on the autosomes could explain the difference in prevalence of PD between males and females at least when considering the current sample size under study. Further studies are warranted to investigate the genetic architecture of PD explained by X and Y chromosomes and further evaluate environmental effects that could potentially contribute to PD etiology in male versus females.

## Introduction

Parkinson’s disease (PD) is an age-related, progressive neurodegenerative disorder. On average males are ∼1.5 times more likely to develop PD compared to females (Moisan et al. 2016). The reasons for the increased risk in males (relative to females) is not well understood. Possible explanations might include different degrees of exposure to environmental risk factors (such as pesticides and heavy metals), putative risk and protective factors (head trauma, caffeine and urate), the influence of sex-specific hormones, differential aging and life expectancy, or potential genetic factors, either linked or independent of these other factors (Nandipati and Litvan 2016; Gao et al. 2016; Ascherio et al. 2004; Taylor, Cook, and Counsell 2007).

There are also differences in the clinical presentation of PD by sex, female patients are more likely to experience dyskinesia and a slower decline in performance of activities of daily living, while it has been shown that male patients have a higher risk of developing cognitive impairment (Iwaki et al. 2020). Symptoms that present into the earliest phases of PD also differ by sex; REM sleep behaviour disorder (RBD) is much more common in males, and depression and anxiety appear to be more common in females (Postuma et al. 2019; Shiba et al. 2000). PD is a genetically complex disease, with a substantial genetic component explained by rare and common variants (Blauwendraat, Nalls, and Singleton 2020). Several large case-control genome-wide association studies (GWAS) have been performed, the most recent of which identified 92 risk signals across 78 loci (Nalls et al. 2019; Foo et al. 2020). Over the last twenty years we have gained a great deal of insight into the genetic architecture and etiology of PD and this now serves as the basis for several therapeutic approaches. However, the interplay between sex and genetics in PD has not been broadly investigated and it is currently unknown whether the genetic risk varies between males and females. Here we investigate whether there is a difference in the genetic architecture of autosomal risk according to genetic sex using multiple large case-control cohorts.

## Methods

### International Parkinson’s Disease Genomics Consortium data

Genotyping data, all derived from Illumina platform based genotyping, was obtained from members of the International Parkinson’s Disease Genomics Consortium (IPDGC), collaborators, and publicly available datasets (Supplementary Table 1). All PD cases were diagnosed using standard UK Brain bank or the MDS criteria (Postuma et al. 2015; Gibb and Lees 1988). Control participants were excluded if they had any known neurological disease. All datasets underwent quality control separately, both on individual-level data and variant-level data, before imputation was performed as previously described (Nalls et al. 2019; Blauwendraat et al. 2019). Quality control steps included: relatedness filtering (PIHAT>0.125, at the level of first cousin one random individual was removed from each pair), removal of genetic ancestry outliers departing 6 standard deviations from the European CEU/TSI HapMap3 populations, removal of samples with call rates <95% and whose genetically determined sex from X-chromosome did not match that from clinical data, as well as samples exhibiting excess heterozygosity estimated by an F-statistic > +/-0.15. The quality control process and underlying scripts for filtering can be found at https://github.com/neurogenetics/GWAS-pipeline. Filtered genotype data was imputed using the Michigan imputation server with the Haplotype Reference Consortium reference panel r1.1 2016 under default settings with phasing using the EAGLE option (Das et al. 2016; McCarthy et al. 2016). For GWAS analyses, variants passing the post-imputation quality criteria of R^2^ > 0.3 and minor allele frequency (MAF) > 1% were included. Data was split into male and female datasets based on genetic sex. Sex specific case-control GWAS were performed using RVTESTS (v20190205) under default settings (Zhan et al. 2016) using logistic regression on genotype dosages adjusted for the following covariates: age at onset for cases and age of last examination for controls (for a small subset age was not available and missing values were imputed with the mean value using --imputeCov), principal components (PC) 1-5 to account for population stratification, and dataset origin. Age was not included in three datasets due to missing data (MF, VANCE) or co-linearity (FINLAND). PCs were calculated from non-imputed genotype data using FlashPCA (v2.0) (Abraham, Qiu, and Inouye 2017).

### UK Biobank data

Imputed UK Biobank (UKB) genotype data (v3) was downloaded (April 2018) under application number 33601 (Sudlow et al. 2015; Bycroft et al. 2018). PD cases were identified using data fields 42032 and 42033. “Proxy” PD cases were included as part of the analyses, considering individuals who reported a parent affected with PD (paternal PD, data field 20107 and maternal PD, data field 20110) since it has been previously shown to share genetic risk with PD cases (Nalls et al. 2019). Controls were set as people with no report of PD and no parent affected with PD and with an age of recruitment over 60 (data field 21022). Covariates were obtained from the data fields: age of recruitment (data field 21022) and Townsend index (data field 189). Individuals were filtered for relatedness (PIHAT>0.125, at the level of first cousin one random individual was removed from each pair) based on the pre-imputed genotype data using GCTA (Yang et al. 2011). Only European ancestry individuals were included from data field 22006. Imputed genotypes were converted to PLINK2. pgen files using PLINK2 (version v2.00a2LM) (Chang et al. 2015) and filtered for missingness (removing samples with variant missingness >0.1 and MAF <0.01), Hardy-Weinberg equilibrium of P ≥1E-6 and imputation quality (R^2^ > 0.8). GWAS was performed using PLINK2 logistic regression with covariates including age of recruitment, Townsend index, and 5 PCs generated by FlashPCA to account for population substructure (Abraham, Qiu, and Inouye 2017). Four GWAS were performed in the UKB data 1) Male PD cases vs male controls, 2) Female PD cases vs female controls, 3) Subjects with a father affected with PD vs controls and 4) Subjects with a mother affected with PD vs controls (Supplementary Table 1). Proxy conversion was performed as previously described (Liu, Erlich, and Pickrell 2017).

### Additional analyses

Post GWAS quality control was applied to remove variants with a MAF <1%, unrealistic beta values for GWAS (>5 or <-5), and multi-allelic variants. Sex-specific meta-analyses were performed using METAL v2018-08-28 under default settings (Willer, Li, and Abecasis 2010). Post GWAS meta-analysis, the following filtering steps were further applied: variants present in at least 13 out of the 19 datasets and displaying an I^2^ heterogeneity value of <80 were included. LD Score regression (LDSC) was performed to calculate the genetic correlation between summary statistics (Bulik-Sullivan et al. 2015).To assess differences in the magnitude of associations between males and females heterogeneity tests performed. Additionally, genetic heritability was calculated excluding the UKB proxies for males and females separately. All figures and statistical calculations were created and performed using R (v4.0.3) or Python (v3.7). The genetic risk score was estimated using IPDGC data, and Nalls et al. 2019 as the reference dataset to define risk-weighted alleles. Locus numbering was obtained as previously described (Grenn et al. 2020). Large effect size variants from the *GBA* and *LRRK2* regions were excluded (rs76763715, rs35749011, rs34637584, rs114138760).

### Data and code availability

All summary statistics are available at https://pdgenetics.org/resources. Six summary statistics tables are made available: 1) Male PD GWAS (all data), 2) Female PD GWAS (all data), 3) Male PD GWAS (no UKB proxy data), 4) Female PD GWAS (no UKB proxy data), 5) Male PD GWAS (no UKB data) and 6) Female PD GWAS (no UKB data). All code used has been made available on GitHub: https://github.com/neurogenetics/Autosomal-sex-differences-PDv2.

## Results

### Initial data overview

After quality control, we included 13,020 male PD cases, 7,936 paternal proxies, 89,660 male controls, 7,947 female PD cases, 5,473 maternal proxies and 90,662 female controls totalling 214,698 individuals (Supplementary Table 1). The odds ratio for PD in males versus females (excluding proxies) was 1.72 (95% CI 1.66-1.78). Meta-analyses at MAF > 1%, variant present at least 13 out of the 19 datasets and displaying an I2 heterogeneity value of <80 resulted in 7,153,507 variants passing quality control for males and 7,141,404 variants for females. No evidence of genomic inflation was observed, with Lambda 1000 and LDSC intercept values of 1.0013 and 0.9548 (SE=0.0077) for male PD GWAS and 1.0008 and 0.9516 (SE=0.0077) for female PD GWAS (Supplementary Figure 1 and 2). In total, 14 and 13 genomic regions reached genome-wide significance in the male and female GWAS meta-analyses respectively, of which 8 were identified in both and 11 were only genome-wide significant in either GWAS. However, all these 11 genomic regions show significant signals at P >1E-4, similar effect sizes and overlapping 95% CIs of regression coefficients (Figure 1). As expected and previously reported in the largest PD GWAS meta-analysis (Nalls et al., 2019), the *SNCA* and *MAPT* loci were the main significant hits in both the male and female specific GWAS (Figure 1), where the top *SNCA* variant was rs356182; (PD_male: P=3.47E-44, beta=0.256, SE=0.0184; PD_female: P=1.41E-25, beta=0.219, SE=0.0209) and top *MAPT* variant was rs75010486; (PD_male: P=1.48E-32, beta=0.244, SE=0.0206; PD_female: P=5.02E-29, beta=0.268, SE=0.0239).

**Figure 1:**
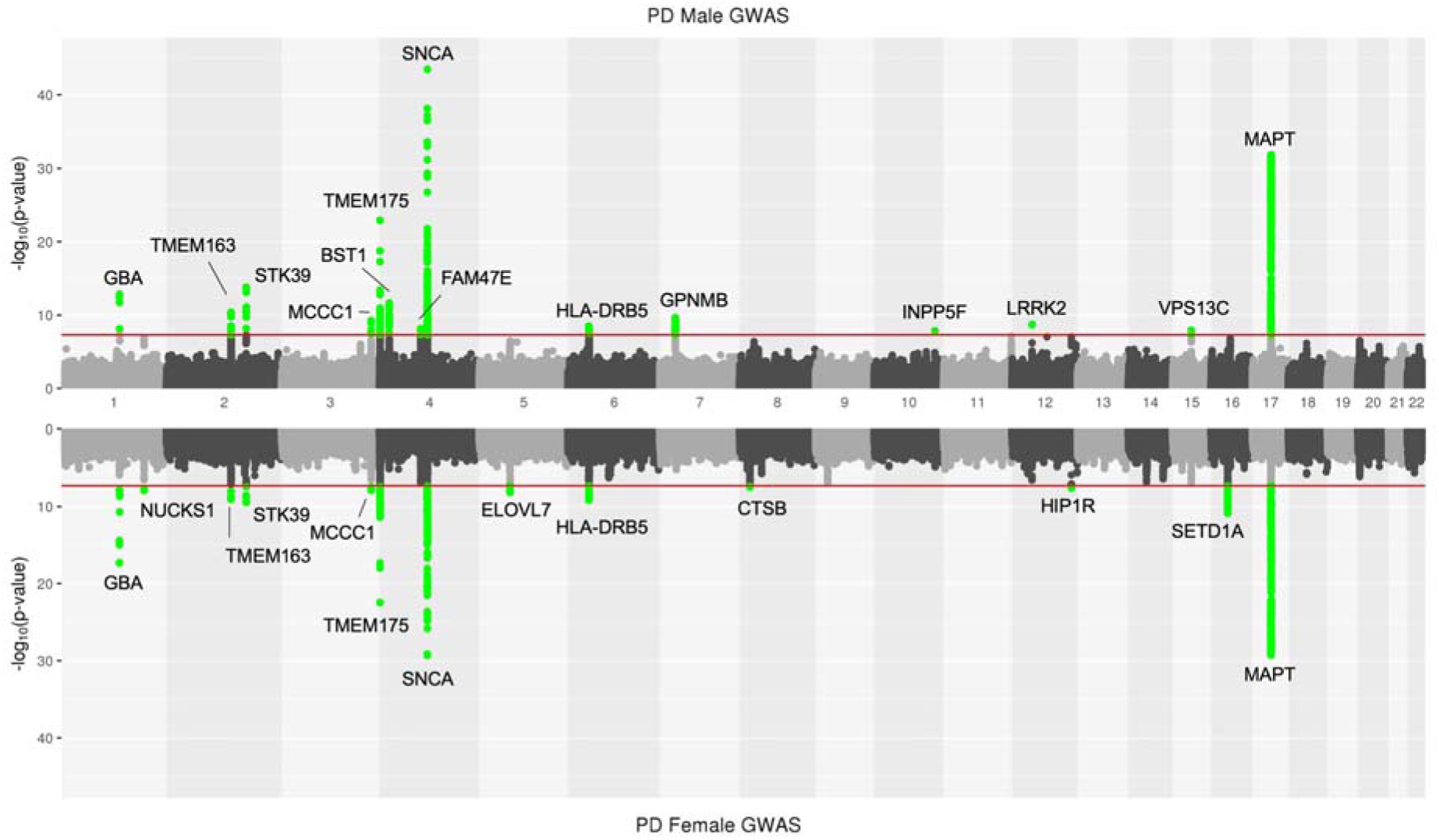
Mirror Manhattan plot of male and female specific Parkinson’s disease GWAS. On the top Male PD GWAS, and bottom Female PD GWAS. Red line indicates the -log10 P-value genome-wide significant threshold of 5E-8. Green dots indicate variants passing genome-wide significance. Signals are annotated based on the closest gene from (Nalls et al. 2019). Figure made using the Hudson Plot Package (Anastasia Lucas, 2020).

**Figure 2:**
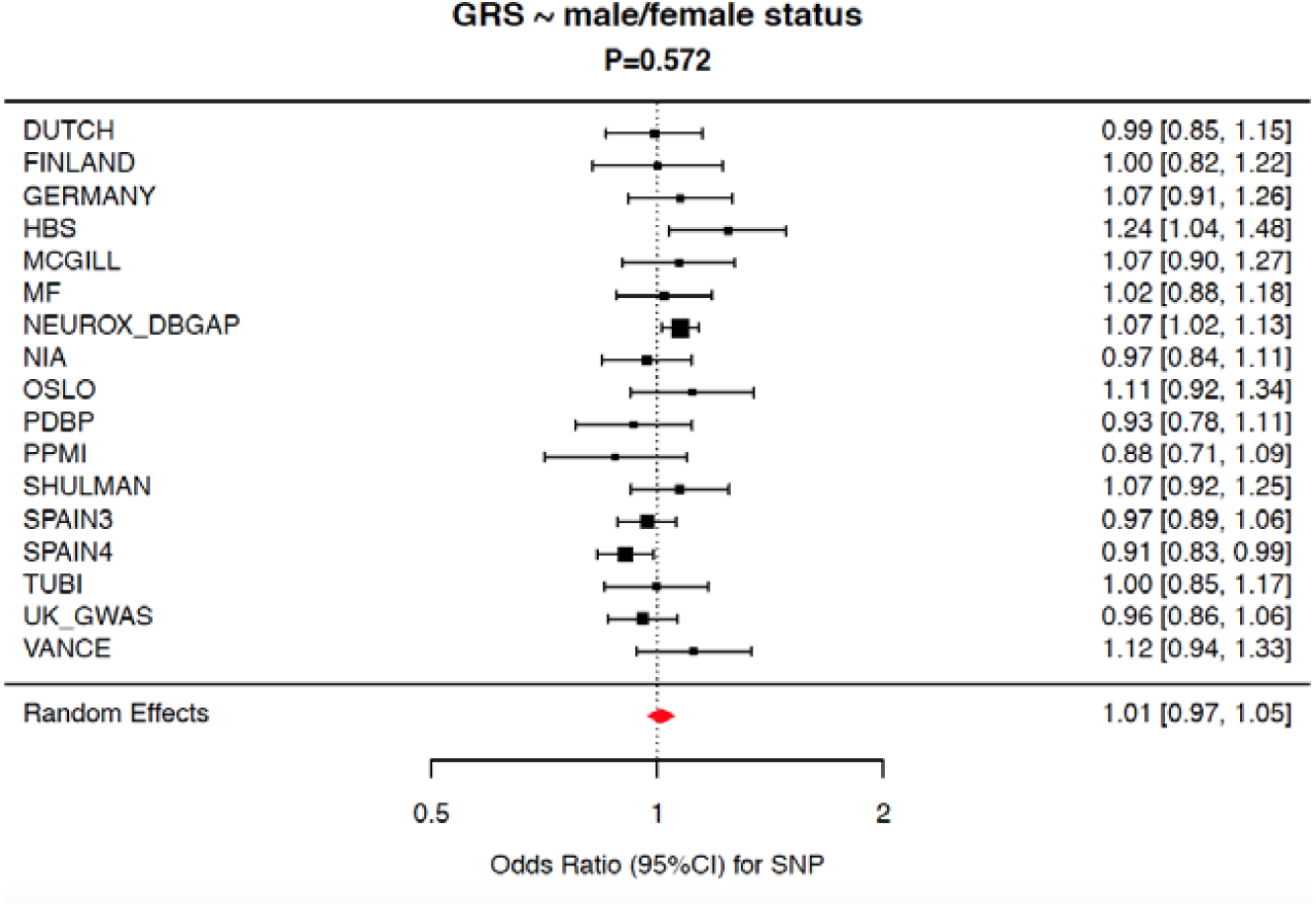
Meta-analysis of the genetic risk score vs male/female status shows no difference between “genetic load” of Parkinson’s disease associated risk. Red diamond indicates the effect estimate (odds ratio) and 95% confidence interval of the aggregate result.

### Comparing Parkinson’s disease risk signals between males and females

To identify potential genetic differences in risk for PD we performed four main analyses, 1) a genome-wide LDSC correlation between male and female specific summary statistics, 2) an analysis of whether the known cumulative genetic risk score was different between males and females, 3) an assessment of whether the known PD risk variants from Nalls et al 2019 were affecting males and females differently and 4) an investigation for potential novel sex-specific assocations in male and female specific summary statistics. The genome-wide correlation using LDSC resulted in a very high genetic correlation between the male and female PD GWASes (rg=0.877, SE=0.0699) which was highly significant at a P=4.24E-36. This shows that on an autosomal genetic level, PD risk is similar between males and females much like how it is similar between self-reported, clinically diagnosed and family history defined cases with genetic correlations at 84% or more as shown in Nalls et al., 2019 when comparing IPDGC (clinically diagnosed) summary statistics with 23andMe data (mostly self-reported) (rg=0.85) or UK Biobank (rg=0.84) (family history defined cases). Heritability estimates (excluding UKB proxies) were similar between males and females (h2_male: 0.2077 (SE=0.0238), h2_female: 0.1857 (SE=0.0304) as expected based on previous estimates.

Next, we tested whether there was a difference of cumulative genetic risk score, based on known GWAS hits and using weights from Nalls et al 2019, between male and female PD cases using the IPDGC data. Each cohort was analyzed separately and results were meta-analyzed. Meta-analysis showed that there was no difference between the cumulative genetic risk score in males and females with PD, P=0.572 (Figure 2). Subsequently, we investigated whether any of the known GWAS risk signals were associated with PD differently between males and females cases. Out of the 90 independent risk signals from the most recent PD GWAS (Nalls et al. 2019), 85 were present in the sex-specific GWAS summary statistics. An almost perfect correlation was observed between the effect sizes of the sex-specific GWASes for these 85 variants (Pearson correlation R2 > 0.95) (Figure 3 and Supplementary Table 2). Similarly, high correlations were found comparing effect sizes between male PD GWAS and Nalls et al. 2019 (>0.98) and females and Nalls et al. 2019 (>0.96). Minor differences were observed in some instances. For example, the *GALC* genomic region (locus 58, rs979812) showed slight differences in magnitude of effect in males versus females; PD_male P=0.06462, beta=0.031, SE=0.0168; PD_female P=3.07E-06, beta=0.0895, SE=0.0192 and P_difference=0.0218. However, none of these were not passing multiple test corrections (Supplementary Table 2).

**Figure 3:**
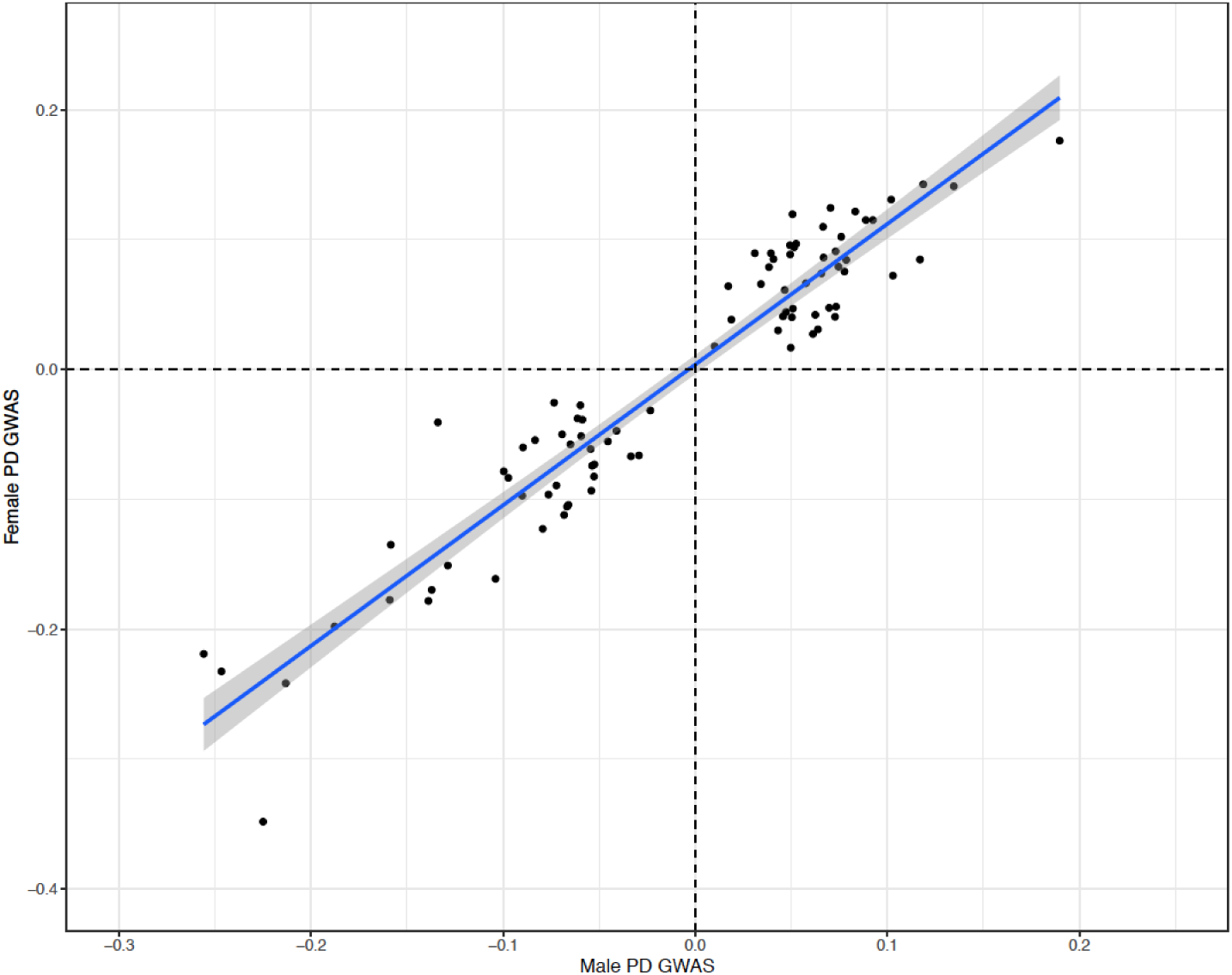
Beta-beta plot of (Nalls et al. 2019) genome wide significant risk signals. Very high correlation (Pearson correlation R2 > 0.95) is observed between effect sizes from the male and female specific GWASes. Additional details can be found in Supplementary Table 2.

Finally, we investigated the full summary statistics to identify potential sex specific hits that have not been identified yet as PD GWAS risk signals. All variants passing P<5.0E-8 from the male PD GWAS were extracted and compared to the summary statistics of the female PD GWAS and vice versa. When plotting the effect sizes of these variants, no differences were observed between male PD GWAS and female PD GWAS effect sizes of variants passing P<5.0E-8 (Supplementary Figure 3 and 4).

## Discussion

Males are ∼1.5x more likely to develop PD compared to females. Here we assessed whether an autosomal genetic difference explains these differences by performing GWAS using several large case-control datasets and separating these by males and females. Overall, based on the results presented here, we could not identify that autosomal genetics contributes to the difference of observed prevalence between males and females. As expected the results from the sex-specific GWASes are highly similar to previous PD GWAS and in particular to the PD GWAS from 2011 with a very similar sample size (15K cases) (International Parkinson’s Disease Genomics Consortium (IPDGC) and Wellcome Trust Case Control Consortium 2 (WTCCC2) 2011).

The question of what actually causes the difference in prevalence will need to be further evaluated. One possibility is genetic variation on the sex chromosomes. Chromosome X contains approximately 850 genes and chromosome Y approximately 70 genes and account for ∼5% and ∼2% of the human genome size respectively. While there is ongoing work in this area, thus far, no association has been found that can explain the significant overrepresentation of male PD patients (Guen et al., n.d.). Given that chromosome Y is only present in males, it could be a good candidate for increased risk. Currently no large genetic association studies have been performed to investigate if certain chromosome Y haplotypes are overrepresented in cases versus controls. There are reports stating that certain Y chromosome genes show male-specific effects for potential dopaminergic loss (Lee et al. 2019). Besides the differences in sex chromosomes in males and females, there are several other not directly assessed here that may likely contribute to sex differences in PD including 1) differences in gene expression in cells and tissues on both autosomes and sex chromosomes (Oliva et al. 2020; Trabzuni et al. 2013), 2) hormone production and 3) the environment including for example smoking behaviour (Heilbron et al., n.d.). All these possibilities need to be studied further with a specific focus on PD.

As for any GWAS study, there are limitations. First, due to the study design, we can only investigate common variants that are present in the imputation panels, meaning that we cannot investigate rare variants and structural variation. Second, given that the majority of the data included in this study was also used in the discovery of the known 90 risk variants from Nalls et al 2019 and sex was used as a covariate in that analysis, it is not suprising that there is a high correlation of effect sizes between the male and female specific GWAS results from Figure 3 due to circularity. However, by using a more unbiased approach in Supplementary Figure 3 and 4 no differences in effect sizes were identified for any genome-wide significant hits from the sex-specific GWASes. Third, although we included a very large number of cases and controls, there could be small effect size variants that play a role in disease, not currently detected due to lack of statistical power.

Overall, here we provide evidence that there are no male or female specific PD GWAS hits and that the difference in prevalence of PD between males and females cannot be explained by common genetics on the autosomes at least when considering the current sample size under study. Further studies are warranted to investigate the genetic architecture of PD explained by the sex chromosomes and further evaluate environmental effects that could potentially contribute to PD etiology in male versus females.

## Data Availability

https://github.com/neurogenetics/Autosomal-sex-differences-PDv2

## Supplementary Tables

**Supplementary Table 1:** Overview of included data

**Supplementary Table 2:** Summary statistics of the genome wide signals from Nalls et al., 2019

## Supplementary Figures

**Supplementary Figure 1:** Quantile-quantile plot of male PD GWAS showing very limited population stratification.

**Supplementary Figure 2:** Quantile-quantile plot of female PD GWAS showing very limited population stratification.

**Supplementary Figure 3:** Effect sizes of the male PD GWAS hits passing genome wide significance plotted versus matching female PD GWAS effect sizes.

**Supplementary Figure 4:** Effect sizes of the female PD GWAS hits passing genome wide significance plotted versus matching male PD GWAS effect sizes.

## Author contributions

Concept and design: CB, MAN, ABS

Statistical analysis: CB, HI, MBM, MAN

Contributed expertise, data or DNA samples:

All Drafting of the manuscript: CB, MAN, ABS, AJN, SBC

Critical revision of the manuscript: All

## Acknowledgements

We would like to thank all of the subjects who donated their time and biological samples to be a part of this study. We also would like to thank all members of the International Parkinson Disease Genomics Consortium (IPDGC). See for a complete overview of members, acknowledgements and funding http://pdgenetics.org/partners. This work was supported in part by the Intramural Research Programs of the National Institute of Neurological Disorders and Stroke (NINDS), the National Institute on Aging (NIA), and the National Institute of Environmental Health Sciences both part of the National Institutes of Health, Department of Health and Human Services; project numbers 1ZIA-NS003154, Z01-AG000949-02 and Z01-ES101986. In addition this work was supported by the Department of Defense (award W81XWH-09-2-0128), and The Michael J Fox Foundation for Parkinson’s Research. This work was supported by National Institutes of Health grants R01NS037167, R01CA141668, P50NS071674, American Parkinson Disease Association (APDA); Barnes Jewish Hospital Foundation; Greater St Louis Chapter of the APDA. The KORA (Cooperative Research in the Region of Augsburg) research platform was started and financed by the Forschungszentrum für Umwelt und Gesundheit, which is funded by the German Federal Ministry of Education, Science, Research, and Technology and by the State of Bavaria. This study was also funded by the German Federal Ministry of Education and Research (BMBF) under the funding code 031A430A, the EU Joint Programme -Neurodegenerative Diseases Research (JPND) project under the aegis of JPND -www.jpnd.eu-through Germany, BMBF, funding code 01ED1406 and iMed -the Helmholtz Initiative on Personalized Medicine. This study is funded by the German National Foundation grant (DFG SH599/6-1) (grant to M.S), Michael J Fox Foundation, and MSA Coalition, USA (to M.S). The French GWAS work was supported by the French National Agency of Research (ANR-08-MNP-012). This study was also funded by France-Parkinson Association, Fondation de France, the French program “Investissements d’avenir” funding (ANR-10-IAIHU-06) and a grant from Assistance Publique-Hôpitaux de Paris (PHRC, AOR-08010) for the French clinical data. This study utilized the high-performance computational capabilities of the Biowulf Linux cluster at the National Institutes of Health, Bethesda, Md. (http://biowulf.nih.gov), and DNA panels, samples, and clinical data from the National Institute of Neurological Disorders and Stroke Human Genetics Resource Center DNA and Cell Line Repository. People who contributed samples are acknowledged in descriptions of every panel on the repository website. We thank the French Parkinson’s Disease Genetics Study Group and the Drug Interaction with genes (DIGPD) study group: Y Agid, M Anheim, F Artaud, A-M Bonnet, C Bonnet, F Bourdain, J-P Brandel, C Brefel-Courbon, M Borg, A Brice, E Broussolle, F Cormier-Dequaire, J-C Corvol, P Damier, B Debilly, B Degos, P Derkinderen, A Destée, A Dürr, F Durif, A Elbaz, D Grabli, A Hartmann, S Klebe, P. Krack, J Kraemmer, S Leder, S Lesage, R Levy, E Lohmann, L Lacomblez, G Mangone, L-L Mariani, A-R Marques, M Martinez, V Mesnage, J Muellner, F Ory-Magne, F Pico, V Planté-Bordeneuve, P Pollak, O Rascol, K Tahiri, F Tison, C Tranchant, E Roze, M Tir, M Vérin, F Viallet, M Vidailhet, A You. We also thank the members of the French 3C Consortium: A Alpérovitch, C Berr, C Tzourio, and P Amouyel for allowing us to use part of the 3C cohort, and D Zelenika for support in generating the genome-wide molecular data. We thank P Tienari (Molecular Neurology Programme, Biomedicum, University of Helsinki), T Peuralinna (Department of Neurology, Helsinki University Central Hospital), L Myllykangas (Folkhalsan Institute of Genetics and Department of Pathology, University of Helsinki), and R Sulkava (Department of Public Health and General Practice Division of Geriatrics, University of Eastern Finland) for the Finnish controls (Vantaa85+ GWAS data). This study was also funded by the Sigrid Juselius Foundation (KM). We used genome-wide association data generated by the Wellcome Trust Case-Control Consortium 2 (WTCCC2) from UK patients with Parkinson’s disease and UK control individuals from the 1958 Birth Cohort and National Blood Service. UK population control data was made available through WTCCC1. This study was supported by the Medical Research Council and Wellcome Trust disease centre (grant WT089698/Z/09/Z to NW, JHa, and ASc). As with previous IPDGC efforts, this study makes use of data generated by the Wellcome Trust Case-Control Consortium. A full list of the investigators who contributed to the generation of the data is available from www.wtccc.org.uk. Funding for the project was provided by the Wellcome Trust under award 076113, 085475 and 090355. This study was also supported by Parkinson’s UK (grants 8047 and J-0804) and the Medical Research Council (G0700943 and G1100643). Sequencing and genotyping done in McGill University was supported by grants from the Michael J. Fox Foundation, the Canadian Consortium on Neurodegeneration in Aging (CCNA), the Canada First Research Excellence Fund (CFREF), awarded to McGill University for the Healthy Brains for Healthy Lives (HBHL) program and Parkinson’s Society Canada. The access to part of the participants at McGill has been made possible thanks to the Quebec Parkinson’s Network (http://rpq-qpn.ca/en/). DNA extraction work that was done in the UK was undertaken at University College London Hospitals, University College London, who received a proportion of funding from the Department of Health’s National Institute for Health Research Biomedical Research Centres funding. This study was supported in part by the Wellcome Trust/Medical Research Council Joint Call in Neurodegeneration award (WT089698) to the Parkinson’s Disease Consortium (UKPDC), whose members are from the UCL Institute of Neurology, University of Sheffield, and the Medical Research Council Protein Phosphorylation Unit at the University of Dundee. We thank the Quebec Parkinson’s Network (http://rpq-qpn.org) and its members. Harvard NeuroDiscovery Biomarker Study (HBS) is a collaboration of HBS investigators and funded through philanthropy and NIH and Non-NIH funding sources. The HBS Investigators have not participated in reviewing the data analysis or content of the manuscript. Data used in the preparation of this article were obtained from the Parkinson’s Progression Markers Initiative (PPMI) database (www.ppmi-info.org/data). For up-to-date information on the study, visit www.ppmi-info.org. PPMI – a public-private partnership – is funded by the Michael J. Fox Foundation for Parkinson’s Research and funding partners, including Abbvie, Allergan, Amathus Therapeutics, Avid Radiopharmaceuticals, Bial Biotech, Biogen, BioLegend, Bristol Myers Squibb, Celgene, Denali, 4D Pharma PLC, GE Healthcare, Genentech, GlaxoSmithKline, Golub Capital, Handl Therapeutics, Insitro, Janssen Neuroscience, Lilly, Lundbeck, Merck, Meso Scale Discovery, Neurocrine, Pfizer, Piramal, Prevail Therapeutics, Roche, Sanofi Genzyme, Servier, Takeda, Teva, UCB, Verily, and Voyager Therapeutics. Parkinson’s Disease Biomarker Program (PDBP) consortium is supported by the National Institute of Neurological Disorders and Stroke (NINDS) at the National Institutes of Health. A full list of PDBP investigators can be found at https://pdbp.ninds.nih.gov/policy. The PDBP Investigators have not participated in reviewing the data analysis or content of the manuscript

