## Supplement for "Investigation of Autosomal Genetic Sex Differences in Parkinson’s disease": Supplementary Figures.docx

Supplementary Figure 1: Quantile-quantile plot of male PD GWAS showing very limited population stratification.

Supplementary Figure 2: Quantile-quantile plot of female PD GWAS showing very limited population stratification.

Supplementary Figure 3: Effect sizes of the male PD GWAS hits passing genome wide significance plotted versus matching female PD GWAS effect sizes.

Supplementary Figure 4: Effect sizes of the female PD GWAS hits passing genome wide significance plotted versus matching male PD GWAS effect sizes.

**Supplementary Figure 1:** Quantile-quantile plot of male PD GWAS showing very limited population stratification. MAF 1% with 7,153,507 variants and Lambda 1000 value of 1.001269 and Lambda = 1.04312.

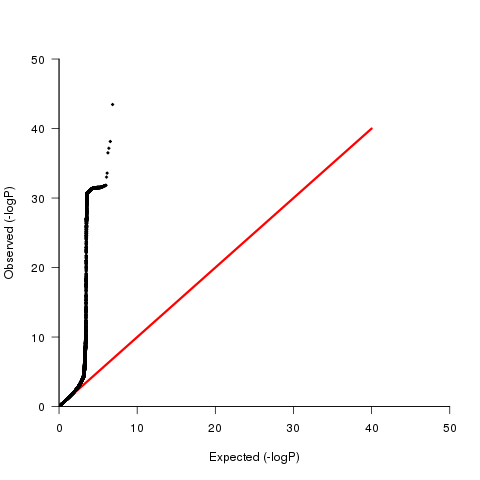

**Supplementary Figure 2:** Quantile-quantile plot of female PD GWAS showing very limited population stratification. MAF 1% with 7,141,404 variants and Lambda 1000 value of 1.0008 and Lambda = 1.019263.

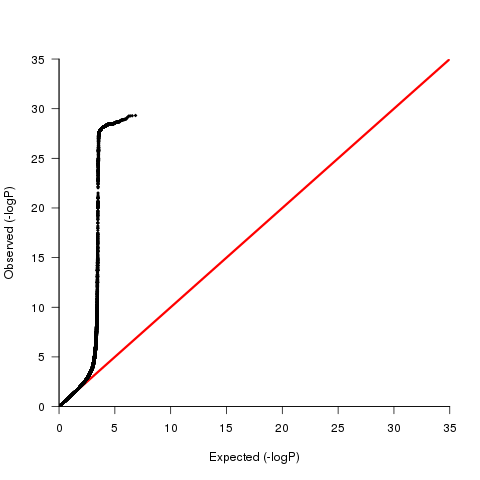

**Supplementary Figure 3:** Effect sizes of the male PD GWAS hits passing genome wide significance plotted versus matching female PD GWAS effect sizes.

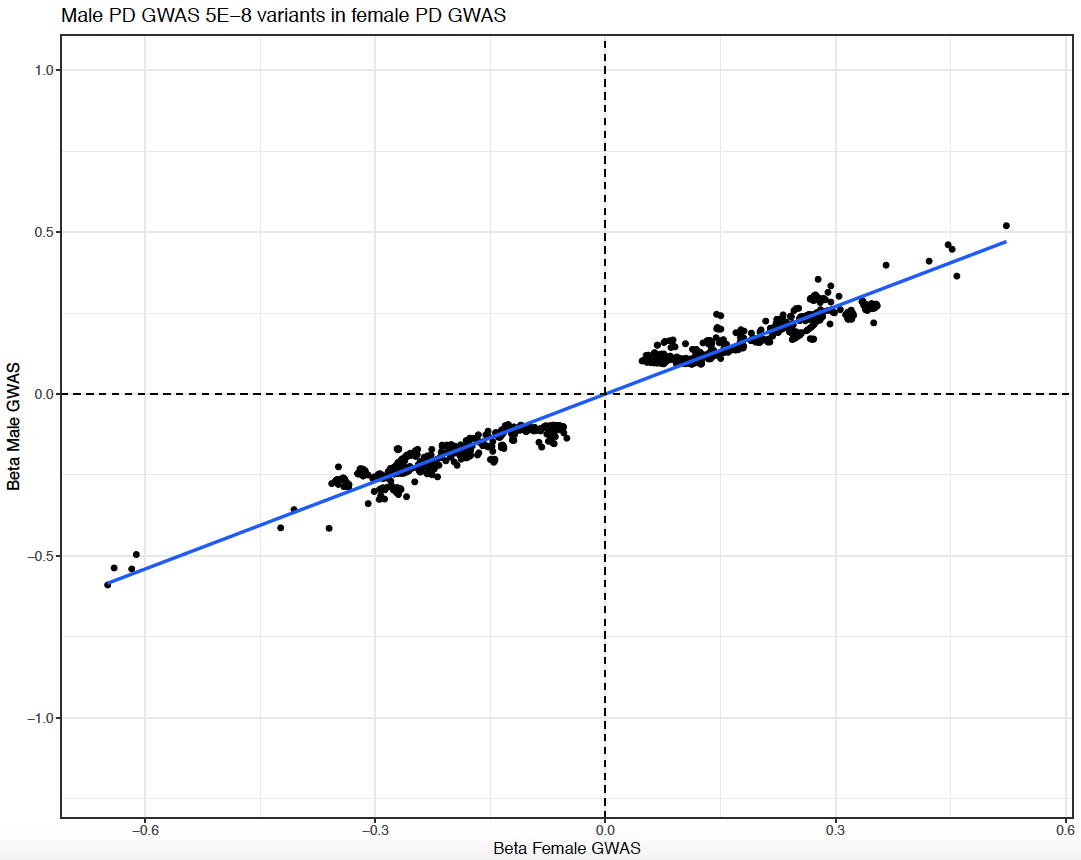

**Supplementary Figure 4:** Effect sizes of the female PD GWAS hits passing genome wide significance plotted versus matching male PD GWAS effect sizes.

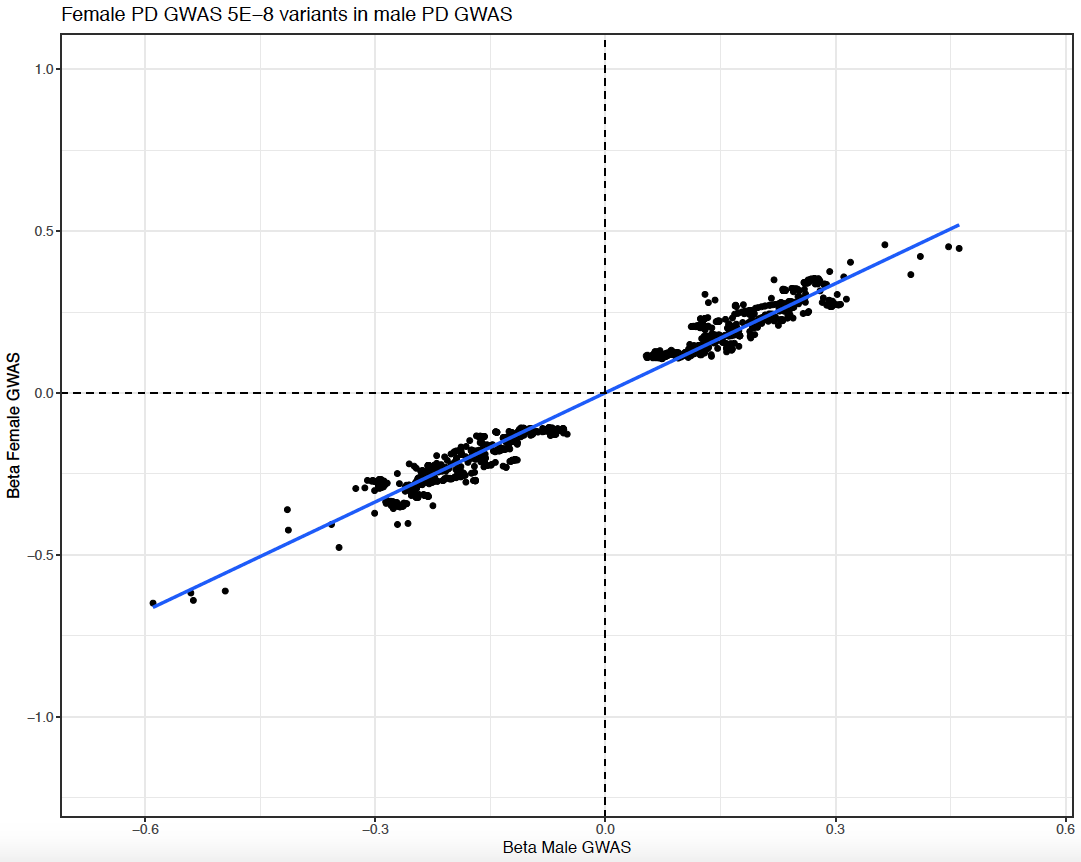
